# A qualitative study of stakeholder perspectives on adopting a digital tool to measure child development at the 2-2½ year review in England

**DOI:** 10.1101/2025.07.01.25330617

**Authors:** JL Lysons, R Mendez Pineda, MRJ Aquino, H Cann, P Fearon, S Kendall, J Kirman, A Lamont, J Woodman

## Abstract

**Aim:** The 2–2½-year review is the final universal mandated contact in England’s Healthy Child Programme, with a child development assessment using the Ages and Stages Questionnaire-3 (AQ-3). Amid a wider digitisation agenda, the UK government is exploring digital alternatives to the paper-based ASQ-3 tool. Understanding stakeholder perspectives is critical for informing implementation.

**Subject and Methods:** 15 focus groups (63 participants), including parents, health visiting professionals, local authority colleagues, and policy officials, analysed using Framework Analysis.

**Results:** Stakeholders reported potential benefits of a digital tool: user experience, service efficiency, and alignment with national digital priorities. However, where services were trying to meet the mandate to review every child aged 2–2½-years with limited resources and workforce, our participants saw a risk that a digital tool might replace a full in-person assessment. Parents and professionals agreed that any digital tool must not compromise the holistic, relational nature of the 2–2½-year review or undermine the universal coverage of in-person contacts with families. Participants highlighted the complexities of digital exclusion, incompatibility with local data systems, and staff training.

**Conclusions:** Digitisation must be implemented carefully to avoid undermining service equity and the core components of the service (universal in-person assessments), must include system interoperability, professional training.

## Introduction

Digital technologies are increasingly shaping public health delivery in England and internationally. In the UK, digital transformation has been at the heart of NHS long-term strategy for several years; the present government’s Plan for Change highlights a shift ‘from analogue to digital’ as one of three key reform shifts to be published in their 2025 10-Year Health Plan (1). Recent government digitisation initiatives highlight ambitions for integrated records, remote monitoring, and wider access for patients; lay out expectations to offer a ‘digital-first’ option for the majority of patients by 2029 (2); for comprehensive electronic health record adoption and enhanced use of digital tools for health campaigns and surveillance across NHS and public health services by 2025 (3,4). Digitisation initiatives are intended to improve service provision by giving patients tools to access information and services directly, reduce administrative burdens on clinicians and staff, and improve NHS productivity and cost efficiency (2).

However, evidence also warns of the potential risks of digitisation, with digital exclusion, particularly amongst vulnerable and structurally marginalised populations, raised as key mechanisms for worsening health inequalities (5–7). Moreover, recent reporting suggests that despite the UK government’s 2025 deadline, 45% of NHS and 47% of central government services still lack a digital pathway (8). Concerns have also arisen about the extent to which digital data are interoperable, with evidence suggesting minimal dataflow from local into national datasets (9,10); resultantly, health data often sit in silos where they cannot be viewed, edited or meaningfully used within or between organisations.

The Healthy Child Programme (HCP) is the public health service for preschool children and their families in England (see Box 1). Based on the theory of proportionate universalism, the HCP has high potential to address health and wellbeing inequalities across the whole population. The HCP, which offers five mandated contacts to all families in the first three years of a child’s life, is currently the focus of UK government policy (1) but is currently facing serious operational challenges (see Box 1). (1) but is currently facing serious operational challenges (see Box 1). The final mandated contact at 2-2 ½ years involves a developmental assessment; since 2015, the tool required for use by the UK government at this review is the Ages and Stages Questionnaire (3^rd^ edition, ASQ-3), a standardised measure that assesses early child development across five key domains (11,12).

The currently licenced version of the ASQ-3 for use in England is a paper questionnaire; however, the shift towards digitisation across the public health sector has resulted in increased calls for a digital tool that can be used to assess child development at the 2-2½ year review (13). Whilst there is a digital version of ASQ-3 available, this would result in additional licensing costs for the Department of Health and Social Care (DHSC) and would not comply with UK data protection law (GDPR) as the licensing company stores data on services in the United States. DHSC recently commissioned a review into alternatives to the ASQ-3 and this work subsequently recommended the Caregiver-Reported Early Development Instruments (CREDI), a free-to-use measure of early child development with digital capacity (13,14). Internationally, initiatives such as the development of the Child Health and Development Interactive System in the USA (15), South Africa’s ‘MomConnect’ programme (16), and Australia’s National Digital Children’s Digital Health Collaborative (17) demonstrate how the use of digital tools have potential to enhance maternal and child health assessment and outcome monitoring programmes.

DHSC has recently commissioned research into the feasibility using the CREDI to measure child development at HCP health and development reviews (18) Understanding stakeholder views on digitising developmental measures of child development for use at the 2-2½ year review will be critical to informing effective policy and practice, and will provide an evidence base for the development of the digital CREDI offering as commissioned by DHSC.

### Research question

What are key stakeholders’ views on the use of a digital tool to measure child development at the 2-2½ year health and development review, including perceived benefits and concerns?

## Methods

Methods are reported in line with the Consolidated criteria for Reporting Qualitative research (COREQ) (19).

### Box 1.

**The HCP and the 2-21/2 year developmental review: key facts**

**Health visiting and the Healthy Child Programme**

Health visiting teams in England lead the Healthy Child Programme 0-5 years. Health visiting is a complex intervention which works with other services to support children’s optimal health and development. All families in England are offered five mandated contacts between 28 weeks of pregnancy and 2-2½ years after birth, which enable the identification of need across the family system, at crucial stages of development (20,21). Families with identified need are offered additional support; this can mean further assessment, the provision of extra information, or signposting to community assets, or more targeted support including additional contacts and onward to referrals to specialist services. This combination of mandated and additional contacts is based on the principle of “proportionate universalism” and is intended to address the full spectrum of health and wellbeing in families, responding proportionately to need (22).

**The 2-2½ year review**

The final mandated contact at 2-2½ years is intended as a wide-ranging, holistic health and development review, covering topics including but not limited to the child’s growth, diet, sleeping, tooth brushing, vaccinations, and weaning (23). This contact also includes a developmental review, where children’s general development including movement, speech, social skills and behaviour, are assessed. Development assessment at this contact has two aims: first, to help detect early developmental delay at the individual level, in order to trigger support pathways where needed (13,24,25); second, to allow for the collection of local-and national-level data for population-level surveillance of child development outcomes across regions and over time. Despite this dual purpose, evidence suggests that most health visiting practitioners are unaware of the population-level monitoring aspect, compounding issues with the flow of developmental assessment data into national datasets for analysis at the local and population levels (24).

**The service delivery context**

Cuts to public health funding have resulted in the reduction of the health visiting workforce by more than 40% since 2015 (26), with workforce issues compounded by rising levels and complexity of need amongst the population (27). In many localities, health visiting services have adapted to these pressures by adopting a ‘skill mix’ model, whereby universal families (i.e. with no additional identified need) may receive contacts from staff nurses (Band 5), nursery nurses (Band 4), or healthcare or public health assistants (Band 2/3) (20,21); families with additional or specialist need are then typically seen by health visitors, who are specialist public health registered nurses (Band 6). Whilst mandated visits should be offered in-person, contacts are increasingly held in a mixture of modalities, including face-to-face at home,in clinic, by telephone, or via a letter from any member of the skill mix team (20,21,28).

HV team members report that the timely completion of the five mandated contacts are seen as markers of a successfully implemented programme and have attained the status of key performance indicators (KPIs) (28). However, health visiting team members stress that rather than being KPI-driven in completion of the ‘task’, the mandated HCP contacts are essential for assessing need amongst all families and for providing preventative support across the whole spectrum of need (28). Health visiting team members have reported that the skill mix model can pose a risk to the efficacy of the health visiting service, as thorough assessment of family need is typically beyond the remit of Bands 2-5 staff (28), and is particularly challenging when a mandated visit is conducted over the phone or via letter (28). This runs the risk of mild-moderate need being missed in universal families that may superficially ‘present well’. Moreover, this model results in fully qualified health visitors handling the vast majority of complex and challenging cases, leading to burnout (27,28). This has further implications for staff retention in a service where 40% of staff are anticipated to leave the workforce within the next five years (27).

### Participants

The present findings constitute part of a larger study commissioned by the DHSC via the Children and Families Policy Unit (CPRU) to meet a rapid policy timeline, investigating the feasibility and performance of available tools for use at the 2-2½ year review (24,29). Our sampling methods and sample characteristics are reported in detail elsewhere (24, Supplementary Material 1). We held 15 focus groups with a total of 63 participants between January and September 2023, to gather their perspectives on the use of a digital tool to measure child development at the 2-2½ year health and developmental review. Seven focus groups were held with parents (*n* participants = 29); five focus groups were held with health visiting professionals (*n* participants = 24) and two with local authority (LA) professionals (*n* participants= 5; clinical service leads *n=* 4, data quality manager *n*= 1), all of whom had experience conducting, managing, or handling data generated by the 2-2½ year review. One focus group was held with English DHSC policy colleagues; participants (*n*= 5) were identified by the study team’s DHSC liaison officer as having a role in early years policy and/or public health nursing.

### Analysis

Focus group topic guides explored participants’ experiences of the 2–2½-year review and their feelings about and priorities for a tool to measure child development for use at the 2–2½-year review, including their perspectives on digitisation of the tool. The sections of each transcript that related to digitisation of the tool were coded and analysed according to the five stages of Framework Analysis (FA) (Figure 1). FA is a qualitative method widely used in social policy and health research (30–32), which allows the systematic categorisation of large volumes of qualitative data, producing highly structured outputs well-suited to dissemination to policy and lay audiences (33). Data were extracted, inductively coded in NVivo V.12, and codes were then used to create an initial coding framework that was applied to the remaining transcripts. The first author (JL) charted data into a framework matrix, which was used to develop themes. Codes, framework, and candidate themes were discussed at regular intervals with members of the study team (JW, RPM).

**Figure 1.**
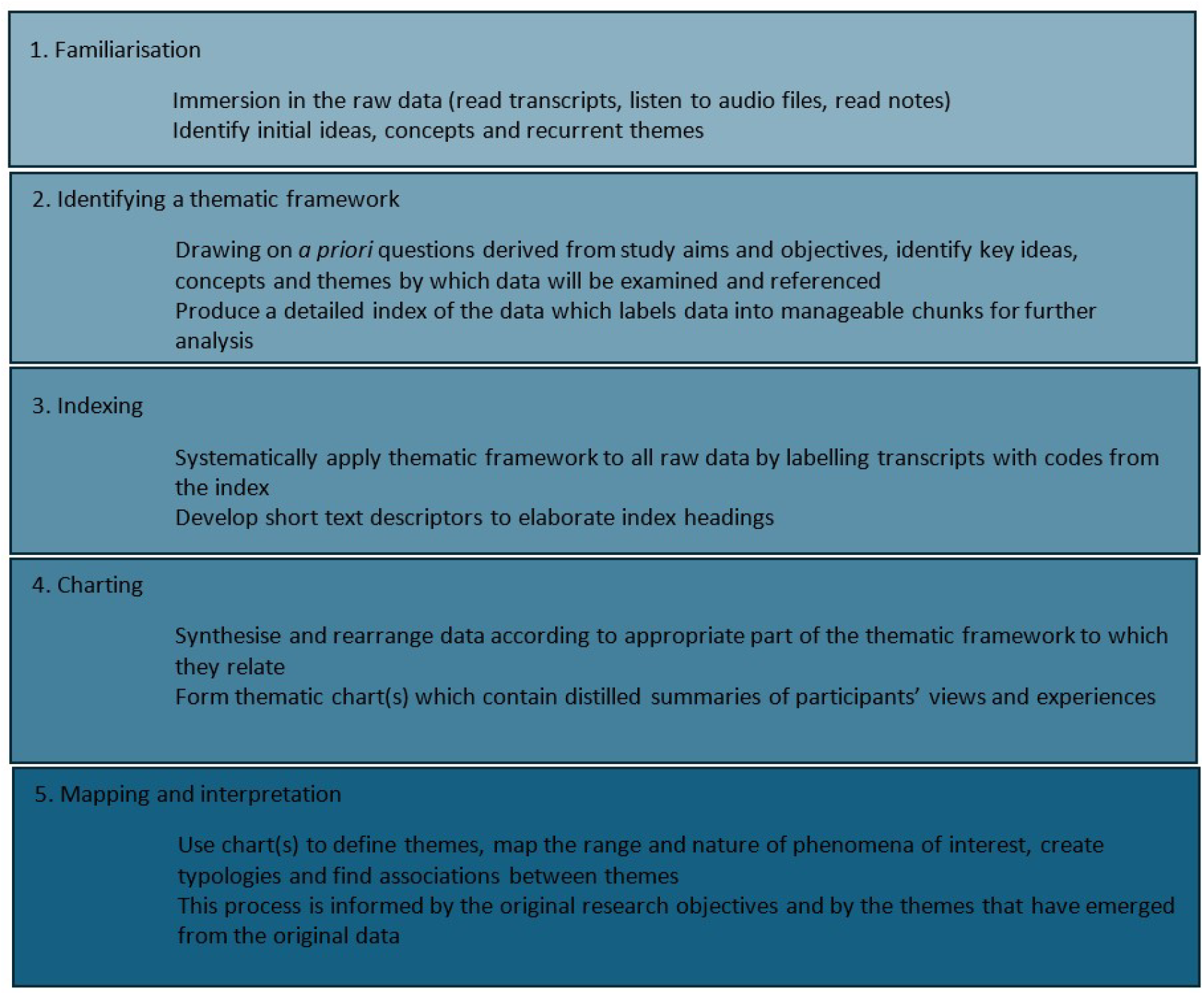
The five stages of Framework Analysis (adapted from Pope, Ziebland & Mays, 2000 (34))

### Ethics

Ethical approval was obtained from UCL Institute of Education and Society Research Ethics Committee (approval number: REC7124, data protection registration number: Z6364106/2022/11/74).

### PPIE

We consulted four parents in a group workshop at the design stage of this study (35).

## Results

Table 1 summarises the perceived benefits of and concerns about the introduction of a digital tool for parents, health visiting professionals, Local Authority (LA) colleagues and policy colleagues at DHSC. Stakeholders perceived the benefits of a digital tool to be: possible improvement of the user experience; modernisation of the system; and improved automation and efficiency. Conversely, stakeholders expressed concerns that a digital tool may, in time, replace valued in-person contacts; may exclude certain groups of people; may be impractical; may be incompatible with pre-existing systems; be costly; and require thorough training and staff motivation.

**Table 1.** Perceived benefits of and concerns about introduction of a digital tool to measure child development.

| PERCEIVED BENEFITS | STAKEHOLDER GROUP |  |  |  |
| --- | --- | --- | --- | --- |
|  | Parents | HV professionals | Policy colleagues | Local authority colleagues |
| **Improving the user experience** |  |  |  |  |
| Constituting part of a centralised digital location for child health and development information | X | X |  | X |
| Improving accessibility by providing translation options and video/aural examples | X | X |  | X |
| Facilitate easier communication and improve relationship between parents and HVs | X | X |  |  |
| **Modernisation** |  |  |  |  |
| Maintaining service reputation and bringing service in line with national digitalisation agenda | X |  | X | X |
| Reduce cost (environmental and monetary) and risk of postage. | X | X | X | X |
| **Automation & efficiency** |  |  |  |  |
| Improve service efficiency by sending scored questionnaires to HV professional before review | X | X | X | X |
| Improve quality of data and dataflow | X |  |  | X |
| Achieve greater reach with less resource |  |  |  | X |
| **CONCERNS** |  |  |  |  |
| **Replacing in-person contacts** |  |  |  |  |
| Slide towards digitisation in under-resourced service could replace valued in-person contacts | X | X |  | X |
| **Universal reach** |  |  |  |  |
| May exclude certain groups e.g. those experiencing digital poverty / areas with poor connectivity | X | X | X | X |
| **Practical difficulties** |  |  |  |  |
| ASQ-3 too long to comfortably use in a digital format | X | X | X |  |
| **Incompatible data management systems** |  |  |  |  |
| Digital tool will need to integrate with plurality of data management systems across country |  | X | X | X |
| **Cost and licencing** |  |  |  |  |
| Negotiating licence for digital tool has potential to be complicated, high-risk and costly |  |  | X | X |
| **Staff training and incentivisation** |  |  |  |  |
| Staff will need training and incentivisation to use digital tool in intended way |  |  | X | X |

### Perceived benefits

#### Improving the user experience

A digital offering was perceived by all groups as being able to improve the user experience by constituting part of a broader, centralised online hub wherein all information about children’s health and development, including access to developmental review materials, would be kept.

LA colleagues emphasised the importance of digitisation efforts to*“really be a system-wide piece of work rather than, you know, a standalone digital ASQ^®^-3”*. Stakeholders suggested that a digital offering may make the tool more accessible by potentially providing text, video, and aural examples of the questions to aid parents’ understanding of child development. They also highlighted the importance of being able to administer the tool in the parent/carer’s primary languages, and hoped that a digital offering might make this a possibility. Parents and health visiting professionals also noted that a digital hub could provide instant messaging channels that would improve parent/carer engagement with the health visiting service, facilitating easier, more personalised communication between parent/carers and the health visiting service.

#### Modernisation: a digital tool for a digital world

> All groups highlighted the need for modernisation of the services. From LA and policy colleagues’ perspectives, a key consideration was the reputational impact of retaining analogue tools/ services; LA and policy colleagues emphasised that providing an online service would fit with government objectives to digitise across all national services. Participants across all groups noted that current reliance on the postal service to distribute key information and materials for the 2-2½ year health and development review is a risky and costly strategy, both in terms of monetary and environmental cost. From a commissioning perspective, LA and policy colleagues at DHSC emphasised that it would be prudent to ensure that any services commissioned in the future are as up-to-date and ‘future-proof’ as possible:
>
> *“I think it’s unrealistic that we can have no digital system for another 10-15 years. It already feels outdated.”* [Policy colleague]

#### Automation and efficiency

A digital tool was perceived as having potential to automate processes, ultimately making the service more efficient. A digital offering was perceived as having potential to save time and money on administration costs, presenting *“an alternative to stuffing envelopes with ASQ^®^-3 questionnaires and posting them out… we’re looking at the capacity of staff it takes to actually send them out.”* [LA colleague].

Parents felt that a digital offering complete with video examples of each ASQ^®^-3 item may help them provide more objective and accurate responses. Policy colleagues at DHSC emphasised the urgent need for data management infrastructure to enable automatic inputting of parents’ ASQ-3 responses into LA systems so they can be *“automatically flowing to the back end for any statistical purposes”* [Policy colleague]. One local authority currently trialling a digital ASQ^®^ -3 confirmed this as a priority as, due to system incompatibility and cost of licencing, their staff had to manually input ASQ^®^ -3 data from the web-based ASQ^®^ -3 data location, to their local data management system, thus effectively doubling the workload.

### Concerns

#### Digital must not replace in-person contacts

Parents, professionals, DHSC policy and LA colleagues took care to emphasise that a digital offering *“should not replace the conversation with the health visitor”* [parent]. A digital option was explicitly framed as something that could enhance, not replace, the 2-2½ year health and development review as detailed above, and that time and energy saved on sending and scoring hard copies of the ASQ^®^ -3 could be spent on other crucial aspects of the review, such as health promotion:

> *“This isn’t a plan to just say ‘just do it online’… even for those families where everything is tickety-boo, there will still be a contact because there are still messages and information to be given at that contact.”* [LA colleague]

However, we also heard that in some places, the digital tool was already being seen as a way to target for the ‘universal’ review. One LA colleague reported that they wanted to *“see how far [we] can stretch the mandation […], not only with skill mix, but actually with digital options, face to face options, questionnaire options”*, indicating that a digital option could replace the in-person health and development review for some people (for explanation of ‘skill mix’, see Box 1).

#### Universal reach

Participants from all groups raised concerns about the extent to which a digital tool may have universal reach. Most commonly, concerns were raised that digital poverty and national inequalities in internet connectivity may exclude certain families. LA and policy colleagues at DHSC acknowledged this and emphasised that *“we wouldn’t ever want a digital-only system so as to be able to include families who are not able to get online.”* [Policy colleague]

#### Practicalities of using a digital tool

Parents and professionals observed that the ASQ^®^-3, particularly in combination with the ASQ^®^ -SE, is very long and so may prove difficult to complete on a laptop/phone in one sitting. A priority for policy colleagues at DHSC was that any digital tool be designed carefully to ensure its fitness for purpose *as “if you’re having to print it out and go in, do it and upload it… that removes the whole purpose of the digital side of it.”* [Policy colleague]

#### Data management and systems

Policy and LA colleagues raised concerns about the need for a digital tool to integrate effectively with local authorities’ current data management systems. Both acknowledged the potential difficulty of devising a digital format compatible with the various data management systems used across the country (e.g., EMIS, Rio, SystmOne). It was important to health visiting professionals that automatically imported parent-reported data be made available in a way *“that we can edit it, rather than it just being what the parent said… that goes on their notes”* [nursery nurse] so that professionals can provide their independent assessment. Health visiting professionals also raised pragmatic concerns that automatically imported ASQ^®^ -3 scores may become out-of-date by the time of their health and development review appointment, as *“quite often parents will rebook their appointments”* [nursery nurse].

#### Cost and licencing

Policy colleagues noted that a lack of a unified data management system could make offering a digital option complicated and costly for LAs. One LA team that had begun the process of negotiating a licence for a digital ASQ^®^ -3 found this complicated and associated with considerable financial risk and ultimately concluded that the risks outweigh the benefits of having a digitally available tool, suggesting that negotiating a digital licence “*should be offered nationally”* [LA colleague].

#### Staff training and motivating practitioners

Policy colleagues at DHSC recognised that “*a lot of training, a lot of guidance, a lot of encouragement”,* and reinforcement of existing infrastructure would be needed to ensure health visiting teams deliver and record the results of a digital tool in the intended way. Similarly, LA colleagues stressed that staff will *“need to be taught and coached”.* A key consideration raised in this group was how the nature of the health visiting role has changed significantly over time, and how a shift to digitalisation would represent further change. Staff buy-in, by providing a clear message on the purpose and correct usage of the digital tool in the context of the health and development reviews, was deemed to be critical:

> *“There are skills with using this kind of technology that they need to adapt to… their course doesn’t really prep them for working in this way. We really need to bring [health visitors] with us, because we can’t lose any more.”* [LA colleague]

## Discussion

### Main finding of this study

We found that parents, practitioners, and LA and policy colleagues cautiously supported digital innovation in the context of the 2-2½ year review but recognised the potential for a digital tool to undermine key aspects of the service, i.e. providing a universal holistic health and development review to all families with opportunities for direct observation of the child and family. These findings align with international evidence that shows successful digital health tools must enhance rather than substitute interpersonal care elements of public health initiatives (36–38). The relational aspect of health visiting is a key component of how health visiting is theorised to work effectively (39,40); our findings confirm that a priority for our stakeholders is that professionals must retain opportunities to observe, engage with, and coach families beyond the use of standardised assessments.

This said, we found that in at least one locality, adoption of a digital tool was viewed as a potentially helpful way to “stretch the mandation”, by using digital-and questionnaire-only options to meet the requirement to review all families with a child aged 2-2½ years (see Box 1). This would pose a significant risk to the HCP’s core function of providing a universal needs assessment to families across the full spectrum of need via in-depth, holistic reviews (28,29). Our findings indicate that any move to digital systems would need significant on-boarding and retraining of staff, both to ensure the appropriate use of digital tool within the context of an in-person holistic health and development review, and to ensure the retention of an already-low workforce.

Many participants saw digital provision being able to improve efficiency by automating time-consuming and often costly processes, particularly by automatically migrating ASQ-3 scores to data management systems. However, practitioners emphasised the importance of ensuring data automation is fully integrated into local systems, enabling access and the ability to edit data to reflect their professional judgement, rather than simply importing parent-reported scores. LA and policy colleagues also highlighted the incompatibility of the different data management systems used across and within regions may inhibit the efficacy of any automation.Interoperability has repeatedly been identified by the UK government as a key barrier to their digitisation agenda (10), with 70% of respondents to a 2024 survey of public sector and government organisations stating that their data landscapes are poorly coordinated and not currently interoperable (8). Evidence from our recent work also shows issues with the flow of developmental assessment data from local to nationally collated datasets, with ASQ-3 data available for only 14% of eligible children in the Community Services Dataset from 2018-21 (9).

Finally, stakeholders described several ways in which a digital offering could improve the user experience, including improving accessibility and facilitating easier communication between parents/carers and the health visiting service. However, stakeholders also observed that digital provision has the potential to exclude certain groups within the population. The risks of digital exclusion for health outcomes are well-documented, particularly amongst low-income and marginalised groups (41–43). Ensuring a twin-track digital and analogue service where parents are, for example, provided with longer appointments and access to the technology needed to complete the ASQ-3 where necessary,will be crucial to avoiding widening existing national health inequalities (10).

### What is already known on this topic

We know that there is a global shift towards leveraging digital technologies in public health and that digital transformation initiatives are often confounded by incompatible systems (3,4,10). In the UK, the government are moving towards digitising non-proprietary tools to measure early child development at the universal health and development reviews, as evidenced by commissioning calls for research into this area (18). We also know that the adoption of digital health tools has potential to exclude certain population groups and to widen

### What this study adds

Our findings demonstrate that digitisation of early child development measures at the 2-2½ year review should enhance, not replace, face-to-face contacts and that, if used to replace mandated contacts for families with no identified concerns, risks missing issues that may only be possible to detect across the wider review. Our findings also demonstrate that a combination of factors, including incompatible data management systems, patchy dataflow and a need to embed the capacity to record professional judgement, could limit the efficacy of a digital offering at the 2-2½ year review. The introduction of any digital provision must be done within a system landscape that is interoperable, capable of effective dataflow, and that is editable by the practitioners on the ground, if it is to make any meaningful improvements to automation or back-end efficiency. Australia’s National Children’s Health Digital Collaborative may provide a useful case study for successful redesign of healthcare service infrastructure with digitisation and system interoperability at its core (17).

### Limitations of this study

For the majority of participants, findings relate to hypothetical rather than actual digital tool usage. Future research should focus on evaluating experiences of localities that have begun trialling digital provision for the HCP health and development reviews. Due to time constraints, we were unable to recruit and employ translators and so focus groups were conducted in English language only. However, small group sizes and experienced facilitators helped ensure conversations were not dominated by single perspectives.

## Conclusion

Digitising measures of child development offers potential to enhance the 2-2½ year review, but must be implemented carefully to avoid undermining the mechanisms by which health visiting and the HCP are understood to work: as a relational practice, whereby practitioners can guide and educate parents, and directly observe and assess the child to promote optimal health and development across the family system. Co-design with practitioners and families will be essential to avoid reinforcing existing inequalities and alienating an already-stretched workforce.

## Data Availability

Data produced in the present study (i.e., focus group transcripts) cannot be made available as stipulated in our ethical approval.

## Acknowledgements

We thank all participants and colleagues who contributed to this study.

## Authors’ contributions

JW, PF and SK conceived the study questions and design. JL, RMP, HC, JK and MRJA conducted the data collection. JL conducted the analysis, with supervision from JW. JL drafted the manuscript and all authors interpreted data, commented on and approved the final manuscript. JW accepts full responsibility for the finished work and the conduct of the study as guarantor, has access to the data and controlled the decision to publish.

